# Real-World Evidence BRIDGE: a tool to connect protocol with code programming

**DOI:** 10.1101/2024.05.08.24306833

**Authors:** A. Cid Royo, R. Elbers, D. Weibel, V. Hoxhaj, Z. Kurkcuoglu, M. Sturkenboom, T. Andres Vaz, CL. Andaur Navarro

**Affiliations:** Department of Data Science and Biostatistics, Julius Center for Health Science and Primary care, University Medical Center of Utrecht, Utrecht University, Utrecht, The Netherlands

**Keywords:** Metadata schema, FAIR, Programming standardization, Electronic Health Records, RWE

## Abstract

**Objective:** *Methods:* Several statistical analysis plans (SAP) from the Vaccine Monitoring Collaboration for Europe (VAC4EU) were analyzed to identify the study design sections and specifications for programming RWE studies based on multi-databases standardized to common data models. We envisioned a metadata schema that transforms the epidemiologist’s knowledge into a machine-readable format. This machine-readable metadata schema must also contain the different study sections, code lists, and time anchoring specified in the SAPs. Further desired attributes are adaptability and user-friendliness.

*Results:* We developed RWE-BRIDGE, a metadata schema with a star-schema model divided into four study design sections with 12 tables: Study Variable Definition with two tables, Cohort Definition with two tables, Post-Exposure Outcome Analysis with one table, and Data Retrieval with seven tables. We provide examples and a step-by-step guide to populate this metadata schema. In addition, we provide a Shiny app that checks the several tables proposed in this metadata strategy. RWE-BRIDGE is available at https://github.com/UMC-Utrecht-RWE/RWE-BRIDGE.

*Discussion:* The RWE-BRIDGE has been designed to support the translation of study design sections from statistical analysis plans into analytical pipelines, facilitating collaboration and transparency between lead researchers and scientific programmers and reducing hard coding and repetition. This metadata schema strategy is flexible by supporting different common data models and programming languages, and it is adaptable to the specific needs of each SAP by adding further tables or fields, if necessary. Modified versions of the RWE-BRIGE have been applied in several RWE studies within the VAC4EU ecosystem.

*Conclusion:* The RWE-BRIDGE offers a systematic approach to detailing what type of variables, time anchoring, and algorithms are required for a specific RWE study. Applying this metadata schema can facilitate the communication between epidemiologists and programmers in a transparent manner.

## 1 Introduction

In the field of pharmacoepidemiology and Real-World Evidence (RWE) generation, retrospective studies on available real-world data are commonplace, focusing on drug utilization, surveillance, and drug and vaccine safety and effectiveness. These studies rely on the rapid analysis of real-world data (RWD), allowing for timely insights into drug usage patterns, adverse events, and the performance of vaccines in diverse dynamic populations. The current state of the art is to work collaboratively with diverse databases using distributed analysis in Europe, America, and Asia. [1] The paper by Gini et al. [2] describes several approaches for multi-database studies. [1, 3, 4, 5]

Extensive collaboration and the broader adoption of RWD and RWE have heightened the demand for standardization, documentation, and transparency of analytical pipelines. Those who conduct studies in the field of pharmacoepidemiology recognize that many decisions are made when creating study variables and analytical datasets, which are often documented inappropriately. Several examples of unreproducible studies are available in the scientific literature, leading to skepticism about the quality of RWE.[6] Initiatives such as RECORD-PE [7], START-RWE [8], and the SPACE framework [9] advocate for more transparency and clarity in the documentation and reporting of RWE studies. Despite the use of Statistical Analysis Plans (SAPs), which include detailed information on these study design and analysis elements, they often lack explicit descriptions to support the development and programming of the analytical pipeline. For instance, plain-language definitions of time anchoring and time windows can lead to overlapping time windows or inaccurate cut-offs in the analytical script. An example is co-variates, which are assessed prior to t0, raising the issue of whether to include or exclude t0. How studies can explicitly and unambiguously document the creation of study variables, time anchoring, algorithms, risk windows, look-back periods, code lists, and rules is still missing. As a consequence, programmers are constantly exposed to making decisions upon these challenges, which require constant support and attention from researchers.

To ease the communication of choices and translation of SAPs, a tool that can concisely and explicitly summarize and document decisions taken during the programming of the analytical pipeline should be developed, ideally in a machine-readable fashion. Although there is abundant literature regarding best practices for conducting and reporting pharmacoepidemiological studies using RWD,[7, 8, 8, 10, 11, 12, 13, 14] there are no concrete methods or tools to translate the information stated in the SAP to the analytical programming pipeline in a machine-readable fashion.

Following the recommendations from the RECORD-PE and REPEAT IT initiatives, we developed a metadata schema that fulfills the following requirements: (1) provides an adaptable and transparent solution to accommodate study specifications from the SAP for the generation of RWE; (2) can automatically incorporate study specifications into the analytical scripts; (3) facilitates communication between collaborators in RWE studies; and (4) adheres to the Findable, Accessible, Interoperable, and Reproducible (FAIR) principles [15]

In this article, we present the RWE-BRIDGE (BRing Intelligence about Data to Generation of Evidence), which, in addition to fulfilling the requirements stated previously, facilitates and improves the documentation of programming decisions with the contribution of both lead researchers and scientific programmers.

## 2 Materials and Methods

This metadata schema was developed for use in the VAC4EU ecosystem, generating RWE on vaccines from RWD in Europe. The VAC4EU ecosystem was built upon the IMI-ADVANCE project, calling for strengthening collaboration in Europe. [16] RWE on vaccines generated within the VAC4EU ecosystem uses the ConcePTION common data model, which is generic and requires only syntactic harmonization. In contrast, semantic harmonization is conducted as part of the study’s analytical script. Further details are described elsewhere [17]

RWD sources originate from different data banks that hold different types of data and have different semantics. [17, 18] Records may be coded with different coding systems or vocabulary like SNOMED CT (Systematized Nomenclature of Medicine – Clinical Terms)[19], ICD (International Statistical Classification of Diseases and Related Health Problems) [20], ATC [21] or even local vocabularies. Depending on the type of vocabulary (hierarchical or non-hierarchical), searches in code lists may be done through exact matching - for non-hierarchical coding systems - or a starting- with approach - for hierarchical coding systems (i.e. hierarchical coding systems are SNOMED CT, ICD10, ICD9, ICPC, and ATC).

The creation of the code lists requires medical knowledge and field expertise; in the VAC4EU ecosystem, code lists are created by a dedicated medically trained group of experts, mapped across terminologies based on the unified medical language system (UMLS) using the Codemapper tool [22]. These code lists are the basis for the semantic harmonization of study variables. For instance, if we have a comprehensive code list containing all codes for each of the supported terminologies, the programmer is able to select the records from the events table in the CDM that are contained in the code list for the specific vocabularies used by the DAP through a simple inner join in SQL:

The VAC4EU ecosystem applies a data engineering and analytical pipeline (hereafter refers as VAC4EU pipeline) where multiple research partners and disciplines work together 1. In general, epidemiologists write the protocol supported by statisticians, and statisticians write the SAP with the epidemiologists. Participating sites (e.g., research partners and Data Access Providers [DAP]) will review those documents. DAPs will extract, transform (step T1), and load their local data into a common data structure (D2). Scientific programmers are responsible for programming steps T2 (creation of study variables and study population) and T3 (application of the design). Additionally, statisticians are responsible for developing the scripts for the estimands (T4), which can be later pooled centrally and post-processed (T5) in tables and graphics for study reports (D6), see figure 1.

**Figure 1:**
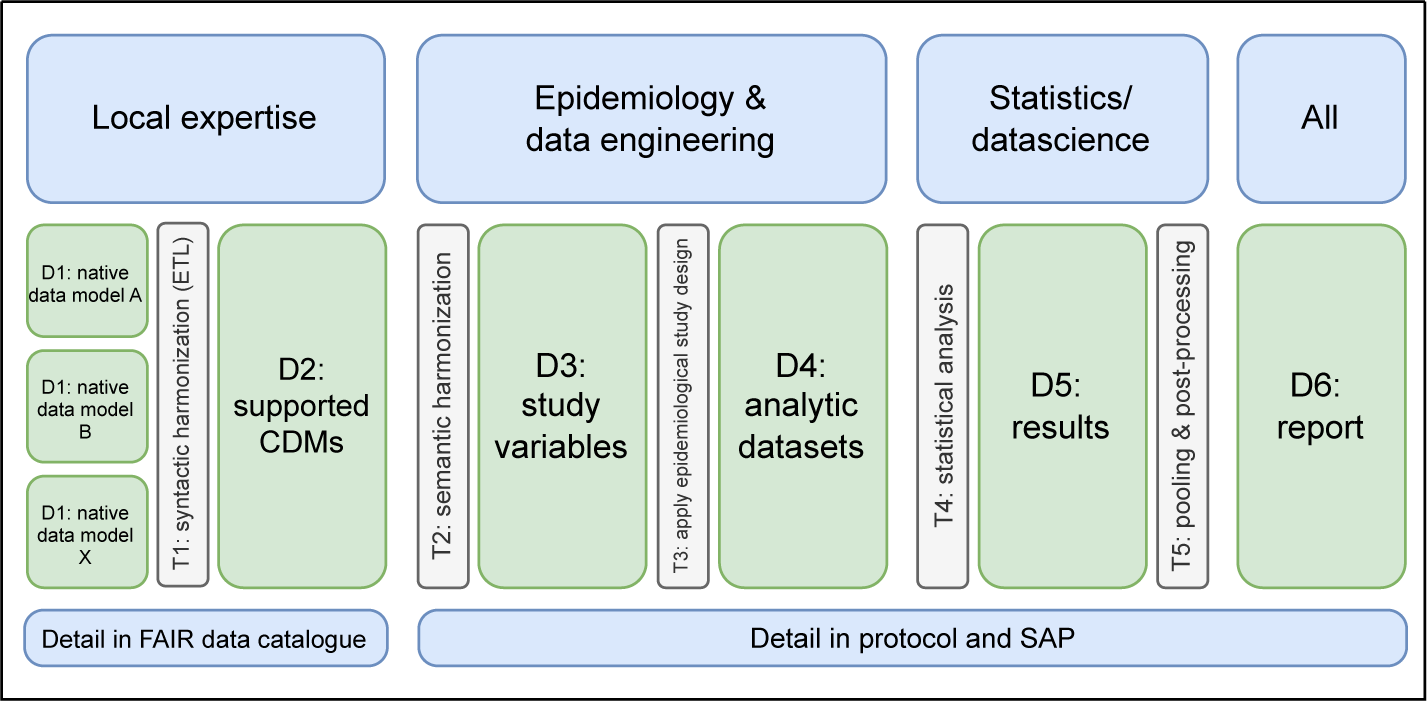
The VAC4EU pipeline is a one-way pipeline that shows all the datasets (D) and data transformation steps (T) applied during a VAC4EU project. The steps are applied consecutively from left to right. The different expertise required in each section of the pipeline can be seen at the top, while the information required for each step and dataset is shown at the bottom. Details on the native data models are necessary at the beginning of the pipeline. These native data models are later extracted, transformed, and loaded (ETL) into the Common Data Model (CDM). Details on the Statistical Analysis Plan (SAP) are needed for step T2 to T5. FAIR: findable, accessible, interoperable, reusable

To design a metadata schema that can transform the relevant SAP content into a machine-readable format, we reviewed multiple SAPs developed within the VAC4EU ecosystem. We identified four main study design elements relevant for RWD engineering in RWE studies: (1) operationalization of study variable (e.g., codes and algorithms) for outcomes, exposure, covariates, and in-exclusion criteria; (2) definition of the population/cohort definition & time anchoring (e.g., observation period, lookback period, follow-up window, censoring); (3) data retrieval; and (4) data analysis. We envisioned each of the main study design elements to be composed of one or more metadata tables, and each table is composed of different fields. In order to identify which tables and which fields are necessary, we have defined a set of questions based on generic vaccines Post Authorisation Safety Studies (PASS) and how these questions can correlate with the metadata schema we proposed; see 4 in 9.1. To improve the understanding of the metadata schema by all partners within an RWE study;[6], we also include fields for descriptions/explanations.

Careful identification of population specifications, medical concepts, or phenotypes to be included, as well as the algorithms and time anchoring in the SAP, is of utmost relevance. Each study variable should be defined in the metadata schema by using either a code list, phenotype, or algorithm, its role in the study (i.e., exposure, covariates, or outcome), and time anchoring. For instance, when defining the look-back period of a covariate, a statement like “A person will be considered immunocompromised when X code is found ever before the index date” may leave us wondering whether the ‘ever before’ definition includes the index date within the time window of consideration. To avoid confusion, it is necessary to explicitly define the start and end of the look-back period with reference to the index date (or t0) as an anchor within our metadata files. This applies to any time window that requires anchoring. In Figure 2 we present an example of two machine-readable metadata files that log window information for the identification of study variables.

**Figure 2:**
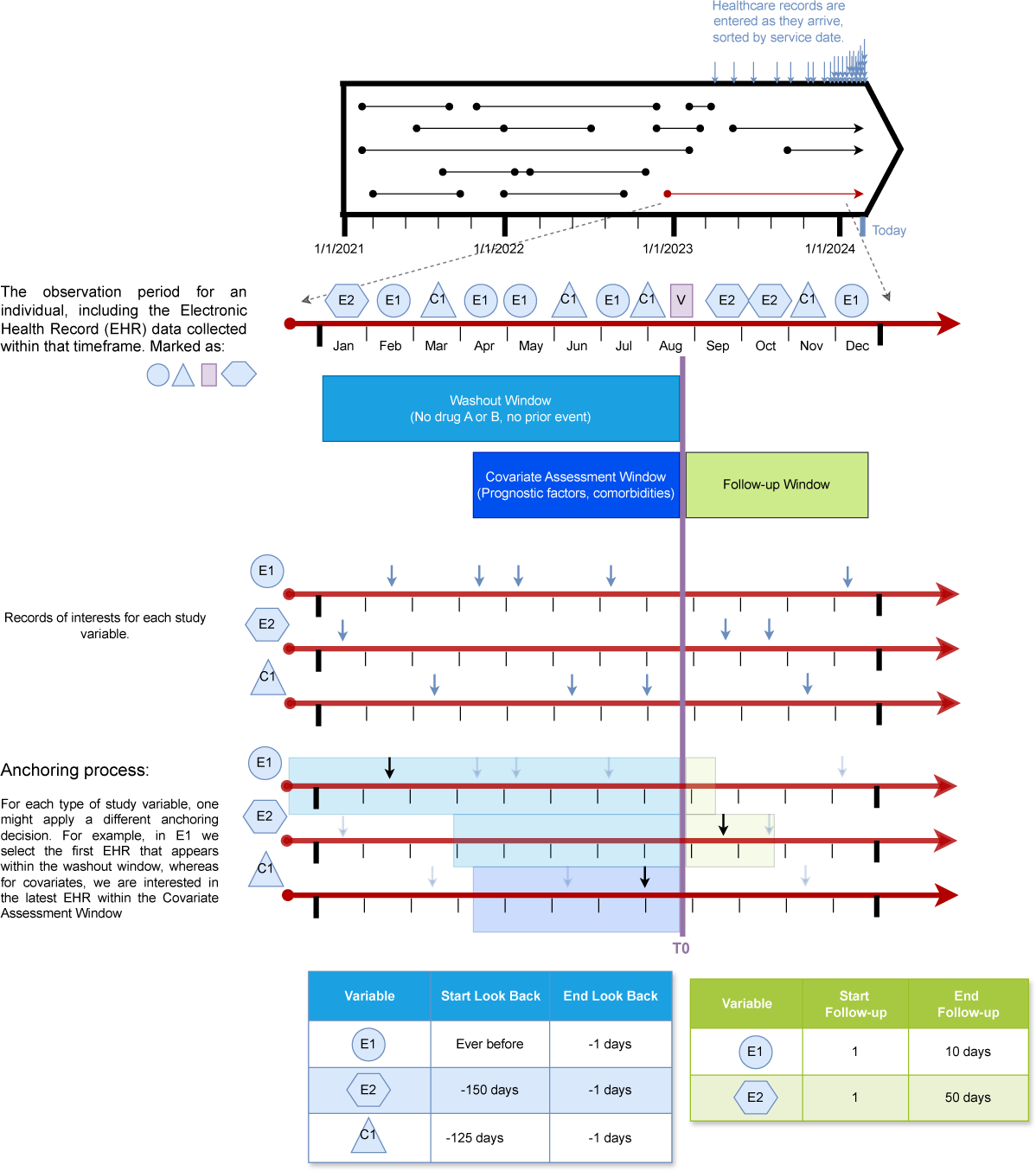
Figure adapted from [23] and expanded with the representation of study variable records and time anchoring process. In the first section of this figure, depicted as horizontal lines, we can see all the observation periods belonging to a person. During observation periods of a person, records are logged into the database. Records are categorized into study variables and then selected for the analytical analysis. The second step is the anchoring process, where T0 is shown in purple, and we set the anchoring window. After the second step, we select a date for the study variable after excluding the records that do not fall within the window. We show two options here: select the earliest or latest records in their respective market, shown in blue. At the bottom, we show how we can define the different characteristics of the time window in a machine-readable format.

The relational database was selected among various database models due to its highly organized and rigid table structure, as well as the flexibility provided by the relationships between tables. Relational databases are powerful and adaptable to various types of data, making them the most suitable choice for our metadata schema. [24] A relational database organizes data in tables, linking them through common fields (foreign keys) for efficient querying. The star schema models a relational database where the central table connects to most secondary tables using foreign keys. In this context, a foreign key is a field in a table linked to another table’s primary key.

## 3 Results

Below, we provide a comprehensive description and a step-by-step guide on manually populating the RWE-BRIDGE metadata schema. For the complete structure, see figure 3. Moreover, we clarify key terms and definitions in table 1.

**Figure 3:**
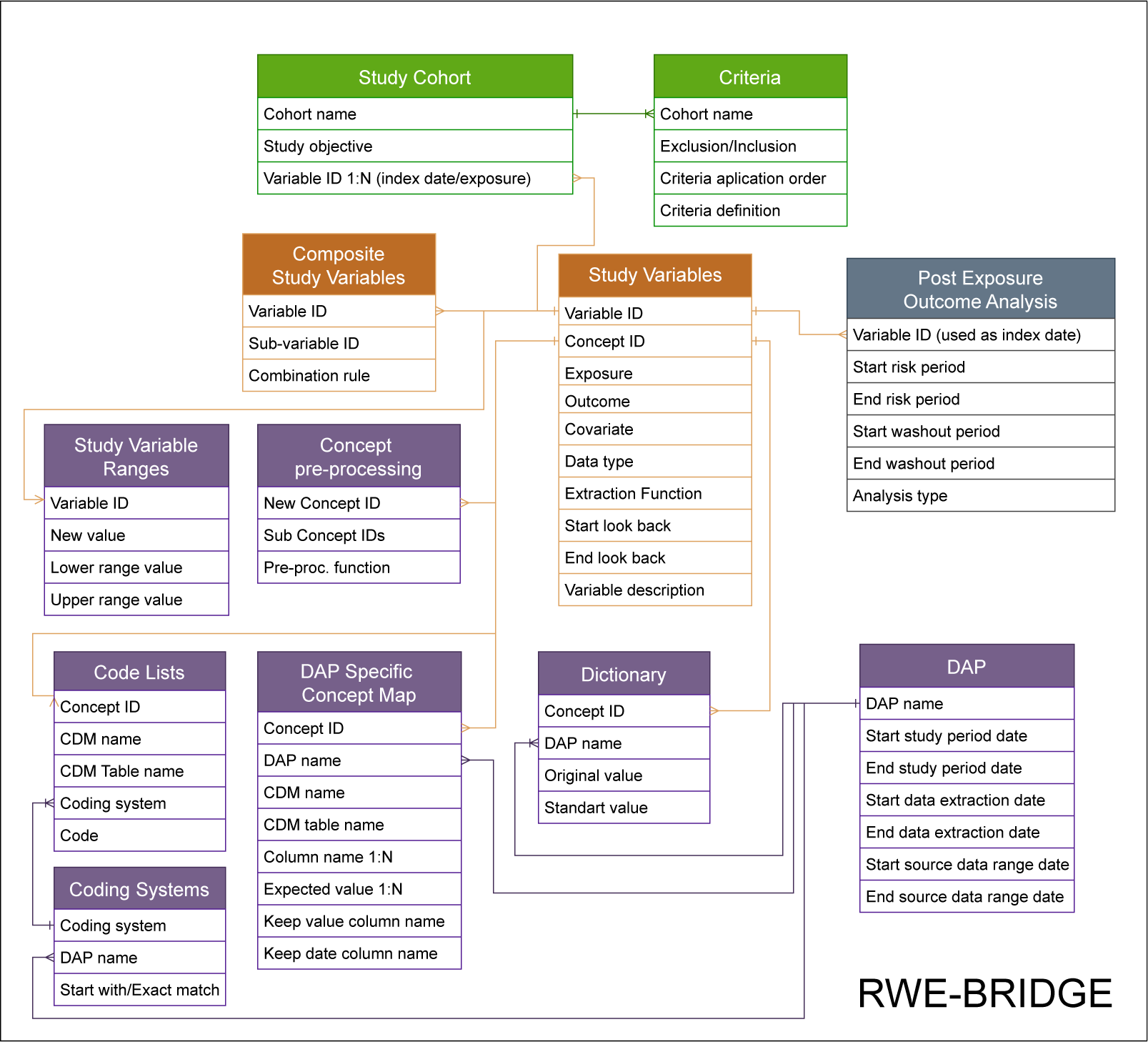
The RWE-BRIDGE diagram. The sections of the metadata schema are color-coded: orange for the variable definition, green for the cohort definition, purple for the data retrieval and processing, and grey for the data analysis.

**Table 1:** Glossary.

|  |
| --- |
| **Field:** Table column. The name of the field is provided in the header. |
| **Foreign key:** Field in a table that refers to unique data values, allowing for the identification of primary key data in another table. These keys connect two or more tables in a relational database. |
| **Medical Concepts and Phenotypes:** Usually, a concept is defined as the collection of clinical codes representing various aspects such as diseases, diagnoses, medications, vaccines, or procedures. The term “phenotype,” as outlined in existing literature, encompasses multiple concepts. |
| **Concept:** In this article, medical concepts, concept sets, and phenotypes are treated as a unified term referring to a concept. |
| **Study Variable:** Person, place, or phenomenon one is trying to measure. In Real-World Evidence (RWE) studies of medicines and vaccines, these typically come from inclusion-exclusion criteria, exposure, outcomes, and covariates. In data engineering, study variables mean selecting/aggregating/arranging records assigned to a person, anchored on time, and selected within a window. |
| **Composite variables:** Combinations of study variables with boolean logic |
| **Anchoring:** Process when assigning a specific date or window of time for a concept - already identified in the study population. For example, the lookback period serves as the boundary for defining the anchoring window between the earliest date and exposure of a covariate or outcome. See Figure 2 for a visual depiction of the anchoring process. |
| **Internal anchoring:** Process when a medical concept is defined based on two or more concepts with a temporal condition between them, e.g., when X happens after Y. |
| **Dynamic population:** Observational datasets are updated constantly through time and stored within a dynamic transactional database, such as real-world data. The population is dynamic, constantly being updated and enriched with new information. Real-world data can be considered |
| **Dynamic transactional database:** A dynamic transactional database is a database capable of handling constant transactions that update, load, and delete records. |
| **Data Extraction Date:** The date when data were extracted from the dynamic transactional database. |
| **Source Data Range:** The source data range represents the calendar time covered by a data source used to create the study population. |
| **Study Period:** The study period defines the calendar time boundaries for data used in creating the analyzed study dataset. This includes information related to exposures, inclusion and exclusion criteria, covariates, outcomes, and follow-up. |

**Table 2:** Example of DAP Specific Concept Map.

| DAP Name | CDM Name | Concept ID | Table Name | Field Name 1 | Field Name 2 | Field Value 1 | Field Value 2 | Field Record Date | Field Record Value |
| --- | --- | --- | --- | --- | --- | --- | --- | --- | --- |
| Example ConcePTION | ConcePTION | BMI | MEDICAL OBSERVATIONS | mo_source_value | mo_unit | bmi | kg/m2 | mo_date | mo_value |

### 3.1 System Description and Structure

Based on the four study design elements identified previously, the RWE-BRIDGE structure consists of the following sections: (1) Variable definition with two tables, (2) Cohort definition with two tables, (3) Data retrieval and pre-processing with seven tables, and (4) Data analysis with one table. These four sections interact through a relational metadata schema, see 3 where all tables are interconnected through foreign keys (Variable ID, Concept ID, Cohort name, DAP name, and Coding system), facilitating the linkage of all elements across tables.

#### 3.1.1 Variable Definition

The section on variable definition consists of two tables: *study variables* and *composite study variables*.

##### Study Variables

The core table of the RWE-BRIDGE metadata schema. This metadata table assigns unique IDs (Variable ID) to each of the study variables that are described in the SAP. The variable ID field in this table serves as a key to connect the *Study Variables* table with the *Composite Study Variables*, *Study Variable Ranges*, and the *Study cohort*. The concept field is used as a key with the *Code lists*, *DAP Specific Concept Map*, and *Concept pre-processing* tables. A medical concept/phenotype should be a list of diagnosis codes and/or integer values and/or categorical values that conceptually define a study variable. A concept is only operationalizable and, therefore, can constitute a study variable when the concept has been anchored to a reference date and to the period of time of interest defined in the Study Variable table with the start and end of the lookback period- with the exception of the exposure variables where the lookback period is not defined. The study variable table, therefore, includes fields that allow specification of the role of the variables and the time windows.

In the *Study Variables* table, one can specify the programming data type (e.g., boolean, categorical, numerical) and which function is used to select records, for instance: the earliest or latest date in the anchored window period or a count of all records - resulting in a numerical study variable - within the anchored window, or a summation of the values within that window (only for numerical variables). In addition, the table includes fields that indicate the variable’s purpose within the study: exposure, outcomes, or covariates. Finally, each variable can be described in plain language in the field variable description, providing further details to support the Variable ID.

##### Composite Study Variables

This metadata table defines an extra level of combination for study variables by using simple logical formulas to combine them into a composite study variable. As depicted in the 3, this table is connected to the *Study Variables* table through the Variable ID. The *Composite Study Variables* table only allows for combining study variables with an OR, AND, and NOT logic. Before creating a composite study variable, the concepts for the study variables to be used should have been anchored, constituting a study variable per se. An example would be the study variable Diabetes Type 1 or 2, which can be expressed using a logic formula of having either Diabetes Type 1 OR Diabetes Type 2 OR using diabetes medication. The RWE-BRIDGE allows the medical concepts of a composite variable to have different lookback periods.

In the *Composite Study Variables* table, the AND and AND NOT logics are only possible when the concepts do not have a condition between each other (i.e., an internal anchoring). For example, a study variable with an internal anchor could be a boolean study variable that categorizes whether a person was hospitalized after a COVID-19 infection. To acquire such a definition, we will identify all those COVID-19 infection records with a hospitalization record within a window of time after their COVID-19 record date (anchor within the study variable). In this case, we must add such a definition as a new concept in the *Concept pre-processing* table.

#### 3.1.2 Cohort definition

The section *Cohort definition* consists of two tables *Study cohort* and *Criteria*:

##### Study Cohort

In this metadata table, the study subpopulations are described by providing names to the cohorts of interest, the study objective, and the start of the follow-up window for the population (t0). This table provides a method to define different populations and the observation and follow-up windows. Additionally, the study objective field allows for a free-text field to explain the use of the study cohort. This table connects to the *Study Variable* table using the Variable ID field as the key.

##### Criteria

This table outlines the in- and exclusion criteria, including the population to which each criterion applies, definitions, and application order. The *Criteria* connects to the *Study Cohort* table using the Cohort name field as the key. The criteria are defined as expressions that can be used directly in the code. This way, the metadata table can be used —following the application order— to automate a sequence of data processing for the study cohort.

#### 3.1.3 Data retrieval and pre-processing

The data retrieval and processing section consists of seven tables:

##### DAP Specific Concept Map

This metadata table describes information on how to gather concepts that are not structured with standardized codes and require combining multiple values stored in different columns or tables within the CDM instance. The *DAP Specific Concept Map* table provides detailed information about each ‘ unstructured’ concept available in the participating DAP. The table also includes the name of the CDM table where the concept records can be found, the name of the column in the CDM table where values are stored, and the columns to keep, which are values and date columns. For example, we want to identify the body mass index (BMI) records for a specific DAP using the ConcePTION CDM,[25] one can refer to the CDM table MEDICAL OBSERVATIONS and ensure that the columns “medical observation meaning” (mo meaning) and the column ‘medical observation unit’ (mo unit) have the values “BMI” and “kg/m2”, respectively. This will identify each patient’s recorded BMI in the observation value column (mo source value) and date (mo date). See table 3.1.3, with the example above.

**Table 3:**
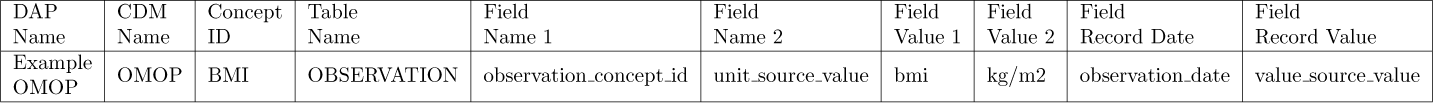
Example of DAP Specific Concept Map for the OMOP CDM.

**Table 4:**
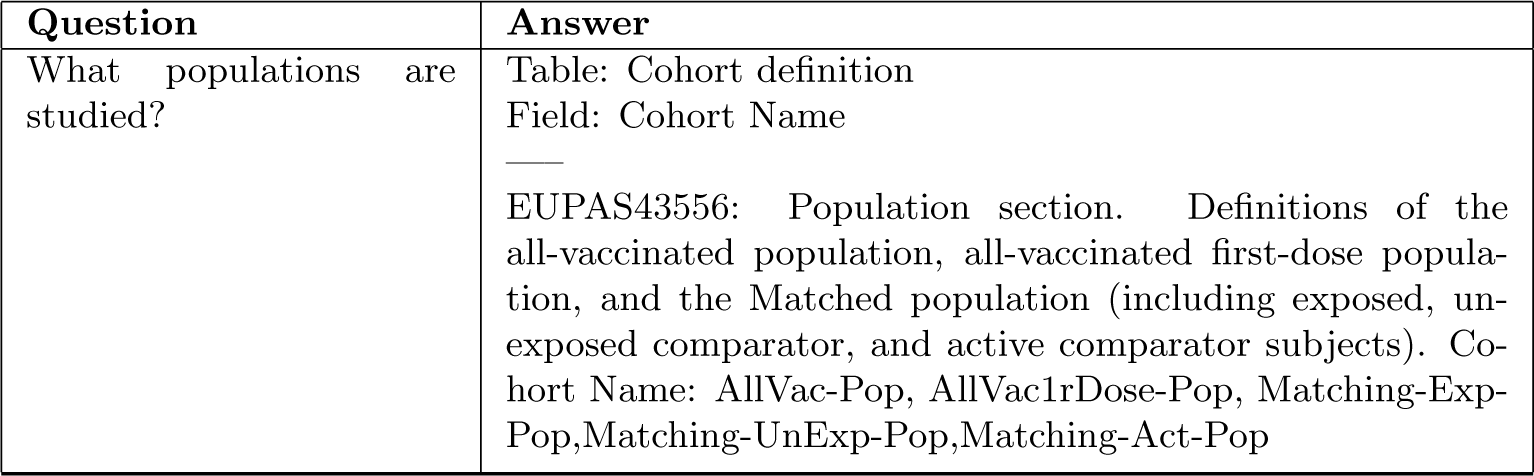
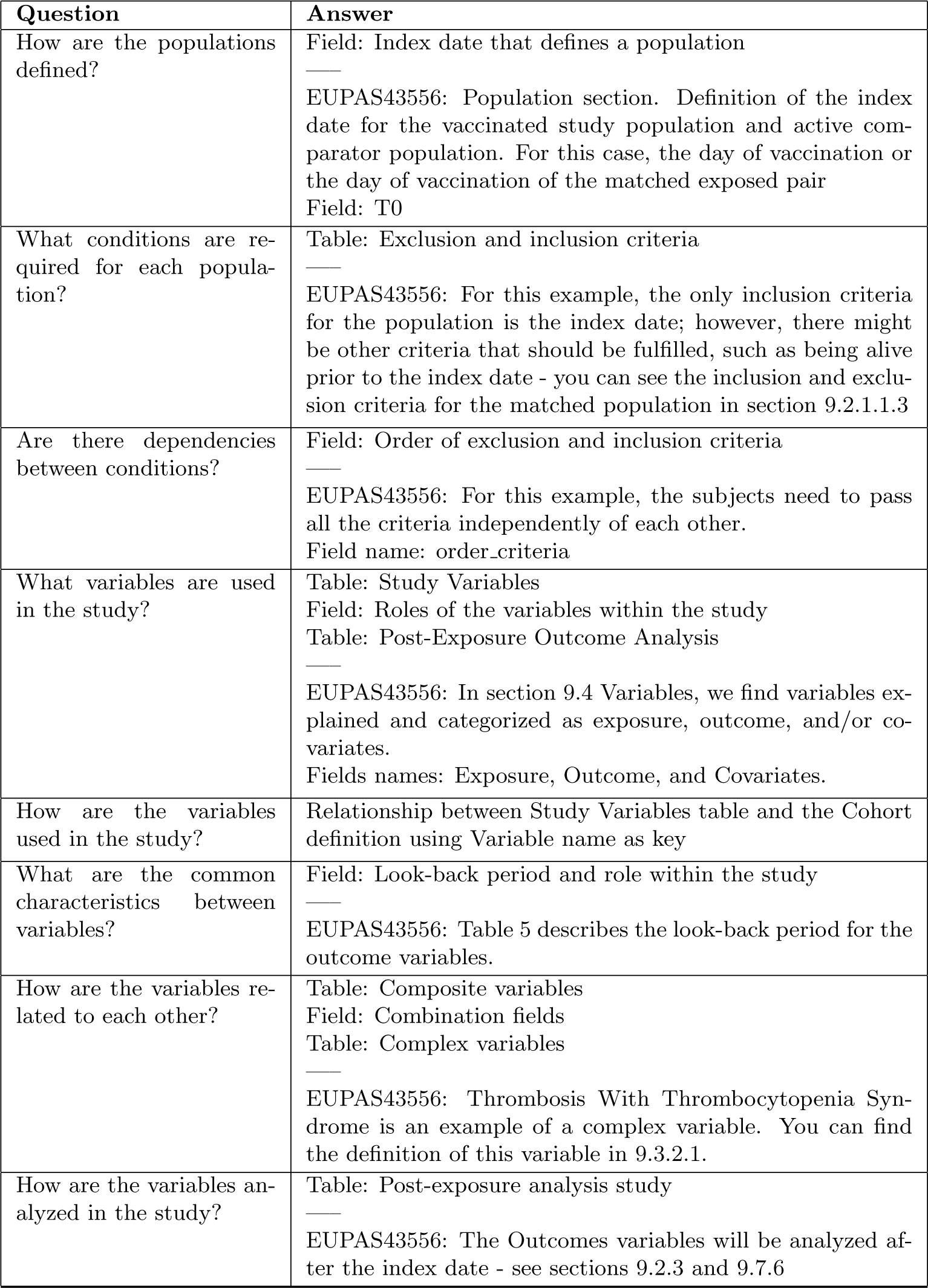
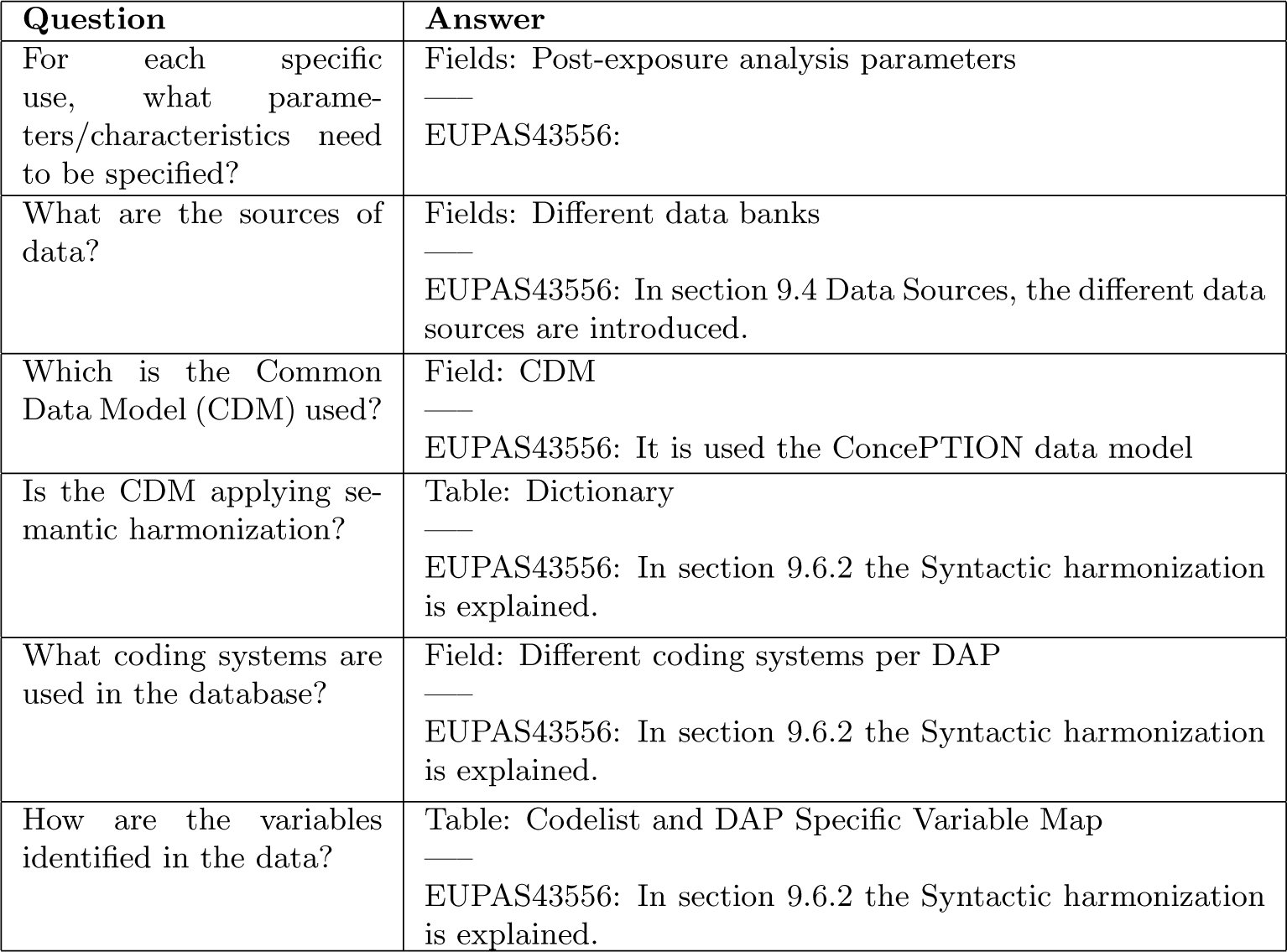
A set of questions is used to identify tables and fields of the metadata schema. We used the study EUPAS43556 updated study protocol as an example. [29].

| Question | Answer |
| --- | --- |
| What populations are studied? | Table: Cohort definition<br>Field: Cohort Name<br>EUPAS43556: Population section. Definitions of the all-vaccinated population, all-vaccinated first-dose population, and the Matched population (including exposed, unexposed comparator, and active comparator subjects). Cohort Name: AllVac-Pop, AllVac1rDose-Pop, Matching-Exp-Pop, Matching-UnExp-Pop, Matching-Act-Pop |
| How are the populations defined? | Field: Index date that defines a population<br>—<br>EUPAS43556: Population section. Definition of the index date for the vaccinated study population and active comparator population. For this case, the day of vaccination or the day of vaccination of the matched exposed pair<br>Field: T0 |
| What conditions are required for each population? | Table: Exclusion and inclusion criteria<br>—<br>EUPAS43556: For this example, the only inclusion criteria for the population is the index date; however, there might be other criteria that should be fulfilled, such as being alive prior to the index date - you can see the inclusion and exclusion criteria for the matched population in section 9.2.1.1.3 |
| Are there dependencies between conditions? | Field: Order of exclusion and inclusion criteria<br>—<br>EUPAS43556: For this example, the subjects need to pass all the criteria independently of each other.<br>Field name: order_criteria |
| What variables are used in the study? | Table: Study Variables<br>Field: Roles of the variables within the study<br>Table: Post-Exposure Outcome Analysis<br>—<br>EUPAS43556: In section 9.4 Variables, we find variables explained and categorized as exposure, outcome, and/or covariates.<br>Fields names: Exposure, Outcome, and Covariates. |
| How are the variables used in the study? | Relationship between Study Variables table and the Cohort definition using Variable name as key |
| What are the common characteristics between variables? | Field: Look-back period and role within the study<br>—<br>EUPAS43556: Table 5 describes the look-back period for the outcome variables. |
| How are the variables related to each other? | Table: Composite variables<br>Field: Combination fields<br>Table: Complex variables<br>—<br>EUPAS43556: Thrombosis With Thrombocytopenia Syndrome is an example of a complex variable. You can find the definition of this variable in 9.3.2.1. |
| How are the variables analyzed in the study? | Table: Post-exposure analysis study<br>—<br>EUPAS43556: The Outcomes variables will be analyzed after the index date - see sections 9.2.3 and 9.7.6 |
| For each specific use, what parameters/characteristics need to be specified? | Fields: Post-exposure analysis parameters<br>—<br>EUPAS43556: |
| What are the sources of data? | Fields: Different data banks<br>—<br>EUPAS43556: In section 9.4 Data Sources, the different data sources are introduced. |
| Which is the Common Data Model (CDM) used? | Field: CDM<br>—<br>EUPAS43556: It is used the ConcePTION data model |
| Is the CDM applying semantic harmonization? | Table: Dictionary<br>—<br>EUPAS43556: In section 9.6.2 the Syntactic harmonization is explained. |
| What coding systems are used in the database? | Field: Different coding systems per DAP<br>—<br>EUPAS43556: In section 9.6.2 the Syntactic harmonization is explained. |
| How are the variables identified in the data? | Table: Codelist and DAP Specific Variable Map<br>—<br>EUPAS43556: In section 9.6.2 the Syntactic harmonization is explained. |

We could then translate the example explained above in an SQL query:

~~~
SELECT person_id, mo_source_value, mo_date
FROM MEDICAL_OBSERVATIONS
WHERE mo_meaning = ‘bmi’ AND
  mo_unit = ‘kg/m2’
~~~

As described in the 3, the *DAP Specific Concept Maps* table connects to the Study Variable table through the Concept ID key field and it also connects to the DAP table through the DAP Name key field.

##### DAP

The DAP ETL’ed metadata table specifies the start and end dates for data extraction, source data range, and study period. Therefore, this table allows the lead researcher to redefine base anchor dates ([12]) if necessary for RWE studies. As we can see in figure 3, this table connects to all tables that contain information pertaining to DAPs; these are *DAP Specific Concept Map*, *Dictionary*, and *Coding Systems*

##### Coding Systems

The table provides information on the coding systems used by each DAP. The key field coding system links it to the Code Lists table.

##### Code lists

Code lists are essential for identifying medical concepts such as symptoms, signs, diagnoses, medicines, vaccines, laboratory tests, and procedures. This table contains the following fields: Concept ID which serves a foreign key that links this table to the *Study Variable* table, coding system to be filled up with vocabularies such as SNOMED CT, ICD10, ICD9, ICPC ATC, the CDM name, CDM table name and the actual codes. Examples of code lists to identify phenotypes can be found in openCodelists.org.[26].

~~~
SELECT EVENTS.person_id, CODE_LIST.concept, EVENTS.code, EVENTS.date
FROM EVENTS
INNER JOIN CODE_LIST
WHERE EVENTS.code = CODE_LIST.code AND
  EVENTS.coding_system = CODE_LIST.coding_system
~~~

If the code list is incomplete—as opposed to the previous example—other approaches should be used to identify the code since every coding system might have different codification approaches (e.g., exact match, hierarchical).

##### Concept pre-processing

This table creates or pre-processes new concepts by processing one or more existing concepts available in the *Study Variable* table. The *Concept pre-processing* table is linked to the *Study Variable* table through the key field *New Concept ID*. This is achieved by referencing programming functions to the pre-processed concept. For instance, the concept of BMI might require pre-processing. In some data banks, BMI may not be available, but height and weight records may be present, which are the components required to calculate BMI, BMI = Weight (kg)/ Height (m). To address this, a function called *create bmi* can be created, which takes both weight and height concepts. Note that this step happens at a concept level. Therefore, it means that the potential function will be applied to all available data within the dataset. The pre-processing function is defined in the *Pre-proc. Function* field in combination with the concepts used - defined in the *Sub-concept ID* field. When filling the pre-processing table, we always add one new row per sub-concept, keeping the New concept ID and the Pre-proc. function the same value.

##### Dictionary

Depending on the CDM used for the RWE study, the *Dictionary* table is required to apply a semantic standardization on the categorical values of study variables. Semantic standardization involves substituting original data values with standardized values specified in a *Dictionary*. Therefore, the content and use of this table will vary for every project due to the different CDM used, different content of the RWE-BRIDGE, and different uses by the programmer. Let’s consider the BMI example mentioned in the *Concept pre-processing* table. This table can be beneficial in scenarios like the one described below: Data instances in DAP1 and DAP2 contain height records. In DAP1, heights are identified by the presence of either ‘cm’ or ‘m’, while in DAP2, they are identified by ‘centimeters’ and ‘meters’. Before calculating the BMI, it’s necessary to convert all height values from centimeters to meters. To do this, we first need to standardize the units of measurement across all values. This is done by using a dictionary to change ‘centimeters’ and ‘meters’ to ‘cm’ and ‘m’, respectively. Once this standardization is complete, we can then apply the transformation to convert ‘cm’ values to ‘m’. The *Dictionary* table can be linked to the Study Variable table through the Concept ID field and to the DAP table through the DAP Name field.

##### Study Variable Ranges

This metadata table is a post-processing table that allows categorizing numerical variables into categories. The table defines the new value to assign and the range values for categorization, and it is used after the anchoring process when the values of the selected records are numerical and can be categorized. The reason why this step is happening after the anchoring and not before it is because there are numerical variables that result from a calculation within the anchoring window (i.e., the total number of medications between the index date and 30 days before it), so the categorization needs to happen after anchoring.

#### 3.1.4 Data Analysis

The Data analysis and processing section consists of one table:

##### Post-Exposure Outcome Analysis

The table for *Post-Exposure Outcome Analysis* outlines the various time frames to consider when conducting an analysis after exposure, such as calculating incidence or prevalence rates. The table includes the start and end dates for risk, control windows, and washout periods fields. [12] Risk windows are defined as the duration of treatment excluding the washout, whereas a washout period is a window of time between other periods; this prevents the undesired carry-over effects between periods of time of a patient). Along with an “analysis type” column to provide further context on each study variable. The *Post-exposure outcome analysis* table is linked to the *Study Variable* table through the key field Variable ID, which is used as an index date for the post-exposure analysis.

### 3.2 Strategy to populate the RWE-BRIDGE

Effective completion of RWE-BRIDGE metadata fields requires close and clear communication between the lead researcher and scientific programmer. The content of the SAP and the specifications of the available data sets are usually intensively and extensively discussed.

#### Step 1: Retrieve information on concepts

The first step to populate the RWE-BRIDGE consists of defining the tables for the *Data retrieval and processing* process (see color purple of figure 4). The DAP is primarily involved in this process because it should provide or make accessible through data catalogs the necessary information to retrieve the variables within the available data instance - which is generally defined during the Extract Transform and Load (ETL) process of CDM. Sometimes, the data retrieval step requires knowledge of the coding systems used in the Electronic Health Record (EHR). A code list of all possible codes used for detecting records categorized in concepts can then be developed using this information.

**Figure 4:**
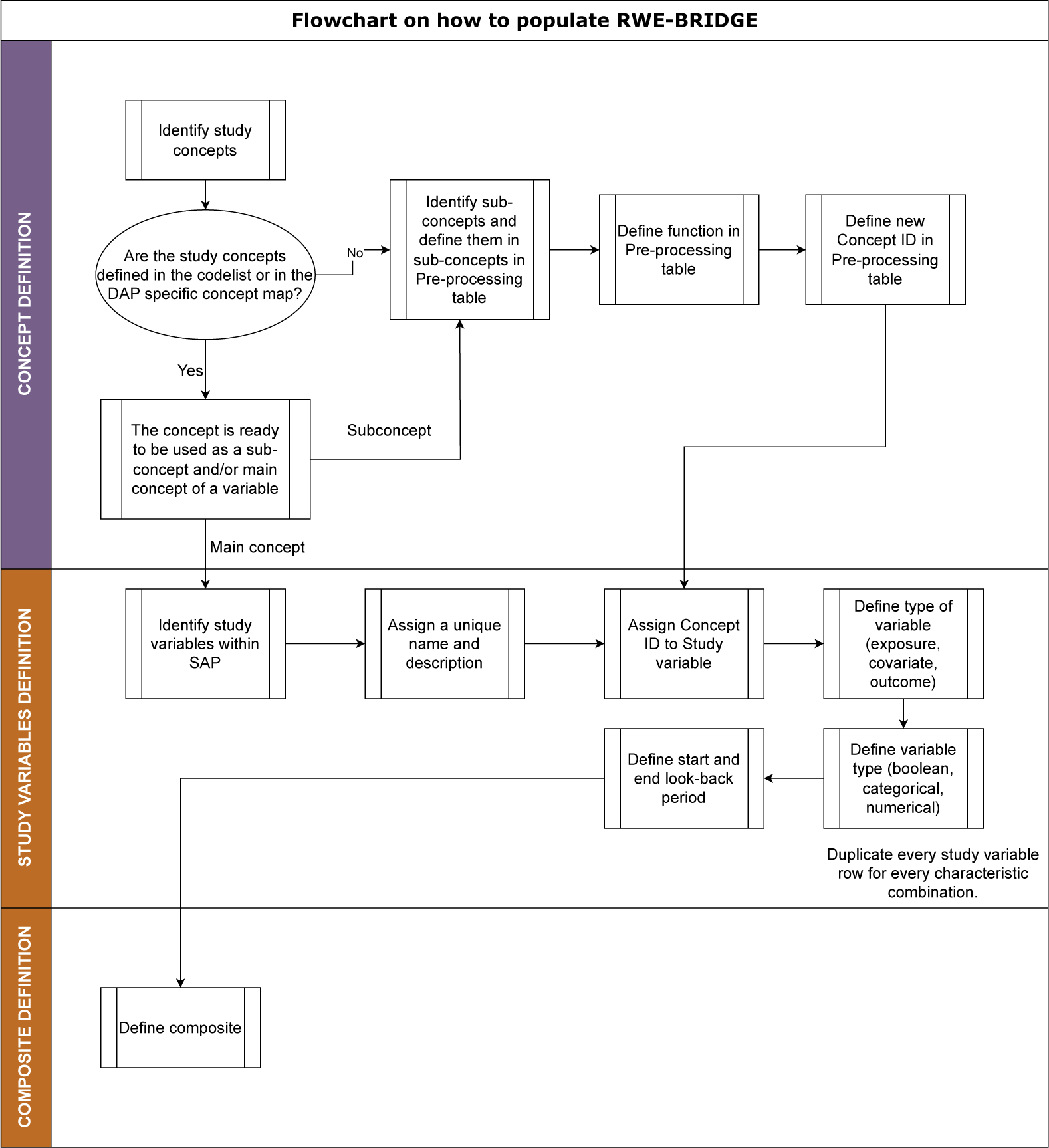
This flowchart outlines the strategy for defining concepts and study variables, starting with concept definition, followed by study variable definition, and ending with the combination of composite variables.

Furthermore, EHR can hold many different types of information and translated into CDM, such as laboratory or BMI values. Therefore, we must provide a flexible solution that can contain the different combinations of values (i.e., either by codes or values) and locations (i.e., CDM tables) required to find any piece of information from the EHR within the CDM of the data instance analyzed. A dictionary should be included in the metadata if semantic standardization is not required in the used CDM. The *Dictionary* should list all possible values found in the data instance for each Data Access Provider and the standard value assigned in the analytical script. For that, one needs to contact the DAP and ask which categorical values are available for each of the categorical variables.

#### Step 2: Build Study Variables

The second step of the strategy is to define the pre-processing of the medical concepts with the *Variable definition process*. In the RWE-BRIDGE logic we begin with the Concept definition followed by the Study Variables metadata file, following the procedure outlined in 4.

#### Step 3: Build Composite Variables and Define Study Variable Ranges

As you can see in figure 4, after completing the Study Variables metadata file, we continue defining Composite Variables. The metadata table for the composite variables outlines the logical formula utilized to combine the different sub-variables. For example, an immunocompromised condition of a person could be defined as the combination of several conditions or diagnosed diseases. Since the different conditions might have different look-back periods, we create a composite variable after anchoring the study variables that compose it. After defining all the study variables - either defined as composite or through previous steps, the *Study Variable Ranges* table can be populated by assigning the category values (either categorical or numerical) to a range of values.

#### Step 4: Creation of study cohort

Defining the *Study cohort process* is the third step, which involves specifying the cohort name and the study objective that the population belongs to. The study variable t0, as outlined in the *Study Cohort* metadata table, will determine the index date of the population. In addition, inclusion and exclusion criteria will be established in the table *Criteria*, which can be described using regular expressions that are easily integrated into the programming without requiring translation.

#### Step 5: Data analysis

The fifth step is defining the *Data analysis*. The *Post-exposure Outcome Analysis* table is available in the RWE-BRIDGE to define outcomes and windows. The proposed metadata schema allows for the precise definition of washout periods and risk windows. These periods are anchored to the index date (t0). The start and end dates for the washout and risk windows’ are defined as the number of days following the index date. For instance, consider a study investigating the risk period for thrombosis following COVID-19 vaccination. Suppose the risk period starts ten days post-vaccination and ends 30 days after the index date. In this case, the time window would be defined as w = [index date + 10, index date + 30]. This approach provides a structured and flexible way to define the timeline of potential effects following exposure.

#### Step 6: DAP dates

This table allows for identifying the primary anchor dates for observation periods that are specified in the *DAP* table, utilizing input from the DAP directly and the study-specific dates.

### 3.3 How to use the RWE-BRIDGE in the analytical script

We designed a flowchart - represented in figure 5 - which outlines the different sequences and steps of using the RWE-BRIDGE in the analysis steo.

**Figure 5:**
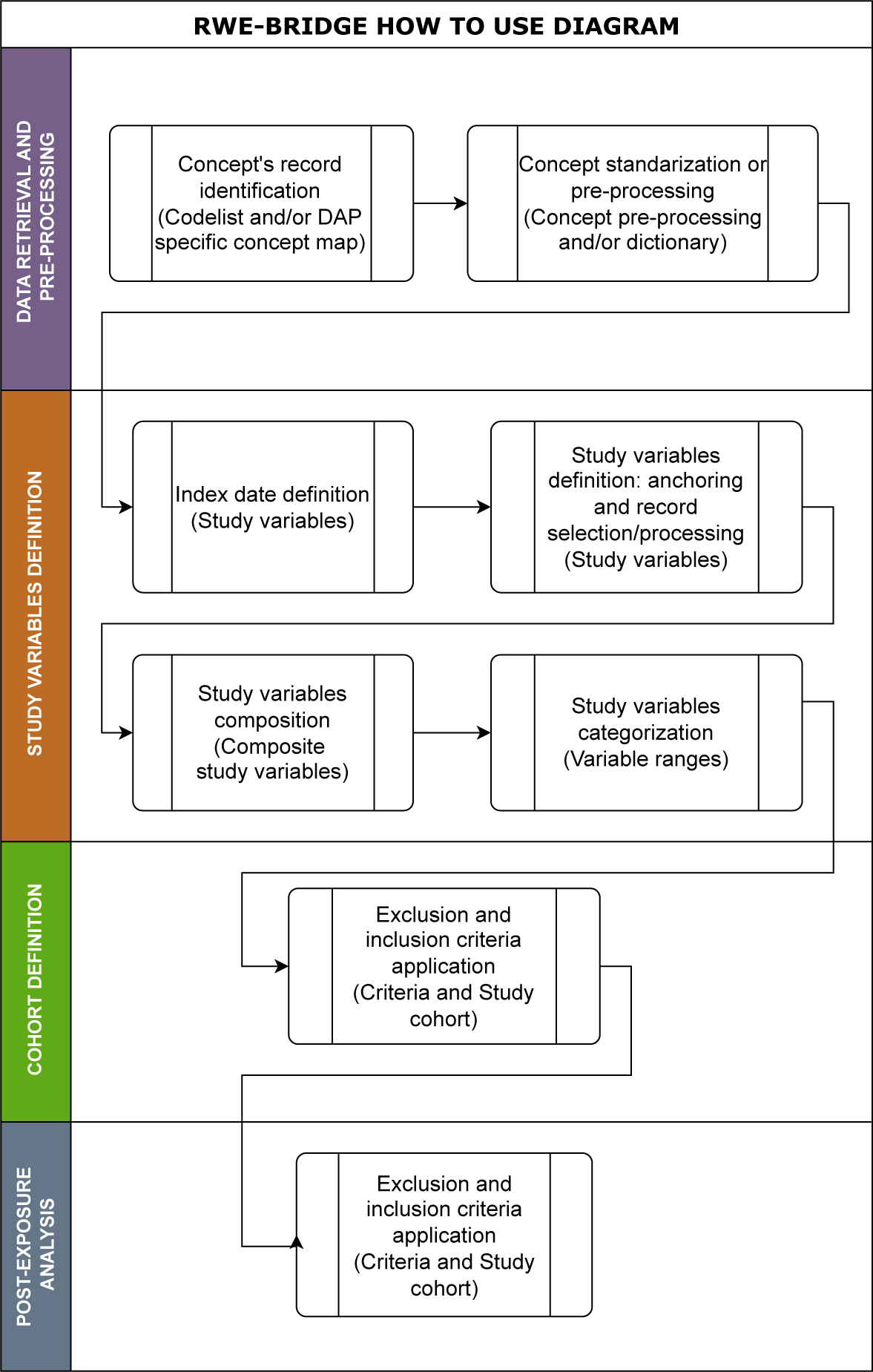
Step-by-step procedure for using the RWE-BRIDGE in analytical scripts.

### 3.4 Metadata check: RWEBRIDGE-CHECKER

An R package has been created with an interactive Shiny App designed to help verify the content and availability of metadata tables in the RWE-BRIDGE schema. Once the RWE-BRIDGE schema has been loaded into the tool, it will display the presence or absence of related tables within the schema. The RWEBRIDGE-CHECKER tool will also ensure that the metadata files in the RWE-BRIDGE schema are consistent.

## 4 Discussion

### Principal findings

In this article, we describe the RWE-BRIDGE, a configuration tool to improve transparency of the analytical programming of RWE studies and communication with epidemiologists.

Based on their complexity, study variables and concepts were categorized into three levels: simple, composite, and pre-processed. Two dedicated tables were created to store the metadata for composite and pre-processed variables. This approach aligns with the phenotype complexity metric (KIP) presented by [27]. According to this metric, the complexity of a phenotype can be defined by three different aspects: Knowledge conversion (K), clause Interpretation (I), and Programming (P). The KIP metric ranges from 0 to 2 per aspect, with 0 being less pre-processed and 2 being more pre-processed. For the RWE-BRIDGE metadata schema definitions, if a variable is defined through a code list, it will have a KIP of (0,0,0). When variables are generated through a DAP-Specific Concepts Map, they will have a KIP of (1,0,0). The composite variables could be (-,1,1) or (-,2,1), where the knowledge conversion (marked with -) would depend on the different study variables that compose a composite. Finally, the pre-processed concepts would be defined as (2,2,2). The three tables within the component of variable definition allow a clear understanding of the complexities of the construction of study variables.

Within the data retrieval and pre-processing section, the *DAP Specific Concept Map* table facilitates semantic harmonization, retrieving unstandardized data (i.e., values) stored across multiple CDM tables within DAPs. Moreover, with the use of the Dictionary table and Concept pre-processing tables, these unstandardized values can be harmonized during the analytical programming. Regarding the *Coding Systems*, *Code list*, and *DAP* tables. Both provide an opportunity to cross-check the required information stated in SAP and the content on the DAP’s ETL data instances; thus, the RWE-BRIGDE also can support the identification of upstream data issues before scripts are run locally within federated analyses.

### Strengths and Limitations

The structure of the RWE-BRIDGE is adaptable, allowing the creation of new tables and fields upon request of specific projects while keeping it as a machine-readable relational database. For example, an additional metadata table that could be useful for a scientific programmer would be to list for each DAP the expected missing concepts; this way, we can differentiate between unavailable concepts and unidentified records for our population - whose aggregated values would be reported as NA and 0, respectively.

The RWE-BRIDGE’s CDM independence ensures CDM interoperability, meaning the metadata schema could be used in multi-database studies with one or more CDMs across the available databases. The DAP-specific variable Map table can prompt the settings for collecting the same variable from different CDMs. For example, the example presented in table 3.1.3 can be edited for the OMOP CDM (see 4). Furthermore, the *dictionary* table allows for standardizing the categorical values we find across different CDMs. The RWE-BRIDGE is compatible with any programming language, as this metadata schema can be structured in a set of different CSV files or within a database (.db) - both these file formats can be loaded and edited by R, Python, SQL, C, and Matlab, among other programming languages.

One limitation is that the manual population of the RWE-BRIDGE can be chal-lenging, requiring knowledge of the SAP and metadata schema. While some fields in the metadata tables may appear unnecessary from a programmer’s perspective, the RWE-BRIDGE serves as a platform for epidemiologists and programmers to convene and deliberate on definitions.

Another limitation of the RWE-BRIDGE is maintaining the consistency of the foreign keys across different tables. To address this limitation and the challenge of the populating procedure, the RWE-BRIDGE checker was developed. This tool can identify any ID not populated in more than one metadata table and detects and reports inconsistencies and missing content within and across metadata tables. The relational structure of the RWE-BRIDGE allows for a systematic validation check of the files’ content, aiding in maintaining data integrity. Details of the RWEBRIDGE-CHECKER can be found in the Additional Materials.

The current design of the RWE-BRIDGE includes only the *Post-Exposure Outcome Analysis* table, this table can be modified to respond to the needs of other study designs or add further tables, if necessary. The current metadata schema includes several tables that were designed to meet the requirements of specific projects. However, it is possible to add or remove tables from the schema to better suit the needs of a particular study.

### Implications for Lead Researchers and Scientific Programmers

The RWE-BRIDGE allows additional metadata tables and fields to create a fully reproducible configuration package design specifically for each RWE study. Nonetheless, the RWE-BRIDGE and its population strategy still have room for development. The template is publically available on GitHub (https://github.com/UMC-Utrecht-RWE/RWE-BRIDGE) and open for comments and improvements.

The RWE-BRIDGE promotes transparency in implementing analytical scripts for RWE generation from RWD. This facilitates reproducibility and assures quality. For example, one can reuse phenotypes that are already available in phenotype libraries. [28] By adhering to the FAIR principles, the RWE-BRIDGE and analytical script can be made publicly available together with a digital object identifier. Furthermore, following the recommendations by [10] and [3], the FAIR characteristics and adaptation of the RWE-BRIDGE create opportunities for applying standard procedures and creating modular data engineering functionality for heterogeneous multi-database studies. Modularization and standardization are key aspects of efficient and reproducible programming.

## 5 Conclusions

Translating SAPs into analytical code is challenging. To overcome this problem, we have developed the RWE-BRIDGE metadata schema to improve communication between epidemiologists and programmers and promote the transparency and FAIRification of RWE.

## Data Availability

No data was used in the present study

## 7 Author Contributions

ACR, RE, VH, TAV, MS, and CLAN conceived and designed the project. ACR, RE, and VH carried out the investigation for the project development. ZK developed the metadata checker R package based on the requirements defined by ACR. TAV and CLAN supervised. ACR led the project and wrote the first draft of this manuscript. All authors revised the manuscript and approved the final version.

## 9 Acknowledgments

Thanks to Rosa Gini and the colleagues from RTI Health Solutions for the engaging discussions about programming challenges within the RWE field.

## 9.1 Additional materials

We have made the template for the RWE-BRIDGE in .csv and .db formats available in the following public repository: https://github.com/UMC-Utrecht-RWE/RWE-BRIDGE. In the following link https://youtu.be/rR1iRSmSCOY?t=1027, a video of the presentation of the RWE-BRIDGE in International Conference of Pharmaco-epidemiology in Halifax (August-2023)

The RWEBRIDGE-CHECKER and its documentation can be found in the following public repository: https://github.com/UMC-Utrecht-RWE/RWEBRIDGE-CHECKER

